# Forskolin-induced swelling of intestinal organoids predicts long-term cystic fibrosis disease progression

**DOI:** 10.1101/2021.02.01.21250609

**Authors:** D. Muilwijk, E. de Poel, P. van Mourik, S.W.F. Suen, A.M. Vonk, J.E. Brunsveld, E. Kruisselbrink, H. Oppelaar, M.C. Hagemeijer, G. Berkers, K.M. de Winter-de Groot, S. Heida-Michel, S.R. Jans, H. van Panhuis, M.M. van der Eerden, R. van der Meer, J. Roukema, E. Dompeling, E.J.M. Weersink, G.H. Koppelman, R. Vries, D.D. Zomer-van Ommen, M.J.C. Eijkemans, C.K. van der Ent, J.M. Beekman

**Affiliations:** Department of Pediatric Respiratory Medicine, Wilhelmina Children’s Hospital, University Medical Center, Utrecht University, 3584 EA Utrecht, The Netherlands; Regenerative Medicine Utrecht, University Medical Center, Utrecht University, 3584 CT Utrecht, The Netherlands; Center for Lysosomal and Metabolic Diseases, Department of Clinical Genetics, Erasmus University Medical Center, 3015 GD, Rotterdam, The Netherlands; Department of Pulmonology, Erasmus MC, University Medical Center, 3015 GD Rotterdam, The Netherlands; Haga Teaching Hospital, 2545 CH The Hague, The Netherlands; Radboud University Medical Center, Radboud Institute for Health Sciences, 6525 XZ Nijmegen, The Netherlands; Maastricht University Medical Center, 6229 HX Maastricht, The Netherlands; Amsterdam University Medical Center, location AMC, 1105 AZ Amsterdam, The Netherlands; University of Groningen, University Medical Center Groningen, Beatrix Children’s Hospital, Department of Pediatric Pulmonology and Pediatric Allergology, Groningen, The Netherlands; University of Groningen, University Medical Center Groningen, Groningen Research Institute for Asthma and COPD (GRIAC), Groningen, The Netherlands; Hubrecht Organoid Technology (HUB), 3584 CM Utrecht, The Netherlands; Dutch Cystic Fibrosis Foundation (NCFS), 3744 MG Baarn, The Netherlands; Department of Data Science and Biostatistics, Julius Center for Health Sciences and Primary Care, University Medical Center, Utrecht University, 3584 EA Utrecht, The Netherlands

## Abstract

Patient-derived organoids hold great potential as predictive biomarker for disease expression or therapeutic response. Here, we used intestinal organoids to estimate individual cystic fibrosis transmembrane conductance regulator (CFTR) function of people with cystic fibrosis, a monogenic life-shortening disease associated with more than 2000 *CFTR* mutations and highly variable disease progression. In vitro CFTR function in CF intestinal organoids of 176 individuals with diverse *CFTR* mutations was quantified by forskolin induced swelling and was strongly associated with longitudinal changes of lung function and development of pancreatic insufficiency, CF-related liver disease and diabetes. This association was not observed when the commonly used biomarker of CFTR function sweat chloride concentration was used. The data strongly exemplifies the value of an organoid-based biomarker in a clinical disease setting and supports the prognostic value of forskolin induced swelling of intestinal organoids, especially for people with CF who have rare *CFTR* genotypes with unclear clinical consequences.

## MAIN

Clinical disease expression in people with CF (pwCF) is variable and results from a combination of genetic, environmental and stochastic factors that are unique for each individual. CF is a recessive, monogenic disease caused by mutations in the cystic fibrosis transmembrane conductance regulator (*CFTR*) gene ^1^. Over 2000 *CFTR* variants which differentially affect CFTR function and clinical phenotype have been identified until now ^2^. The more common mutations have been categorized into distinct classes according to the mechanism by which CFTR function is disrupted ^3^. To better understand how CFTR function contributes to disease expression, biomarkers such as sweat chloride concentration (SCC), intestinal current measurements (ICM) and nasal potential difference (NPD) are used to estimate individual CFTR function. These biomarkers have mostly been validated in the context of CF diagnosis, but their ability to accurately discriminate between pwCF with differential disease progression is limited despite clear relations at population level ^4–10^. Forskolin induced swelling (FIS) of patient-derived intestinal organoids is an in vitro biomarker that quantifies CFTR-dependent fluid transport into the organoid lumen ^11,12^ and may provide a more precise and accurate estimation of CFTR function compared to other biomarkers. Small proof of concept studies showed that FIS correlates with SCC and ICM and that clinical disease phenotypes could be stratified based on FIS level ^13,14^. We hypothesized that individual CFTR function measured by FIS could enable the prediction of long-term disease progression defined by rate of FEV1pp decline and development of co-morbidities such as pancreatic insufficiency (PI), CF-related liver disease (CFRLD) and CF-related diabetes (CFRD). Predicting long-term disease progression provides important prognostic insight, which is especially relevant to people with rare, uncharacterized *CFTR* genotypes or *CFTR* genotypes with varying clinical consequences.

In this retrospective cohort study, we collected data from 176 participants with CF of whom intestinal organoid measurements were available and who provided informed consent to retrieve clinical data from the Dutch CF Registry via the Dutch CF Foundation. Three participants were excluded from the analysis because clinical data was not available, no data was excluded based on organoid measurements. Participant characteristics and individual genotypes with corresponding mutation classification and combination of mutation types (mutation group) are listed in supplementary table 1 and 2, respectively.

Individual FIS responses of all participants after 1 hour stimulation with different forskolin concentrations are shown in figure 1a. Between-subject variability was most apparent at 0.8 μM and 5.0 μM forskolin, but no evident clustering was observed. Consistent with prior studies investigating relations between FIS and CF disease or biomarkers ^12,13,15^, our analyses were performed with FIS levels upon 0.8 μM forskolin stimulation. FIS at 0.8 μM forskolin highly varied among participants (median AUC 142.5, IQR 30.3-1214.9; figure 1a and supplementary figure 1a) as well as within genotype classes (figure 1b-c) and between genotype groups, as defined by the combination of the two mutation types (supplementary figure 1b).

**Figure 1:**
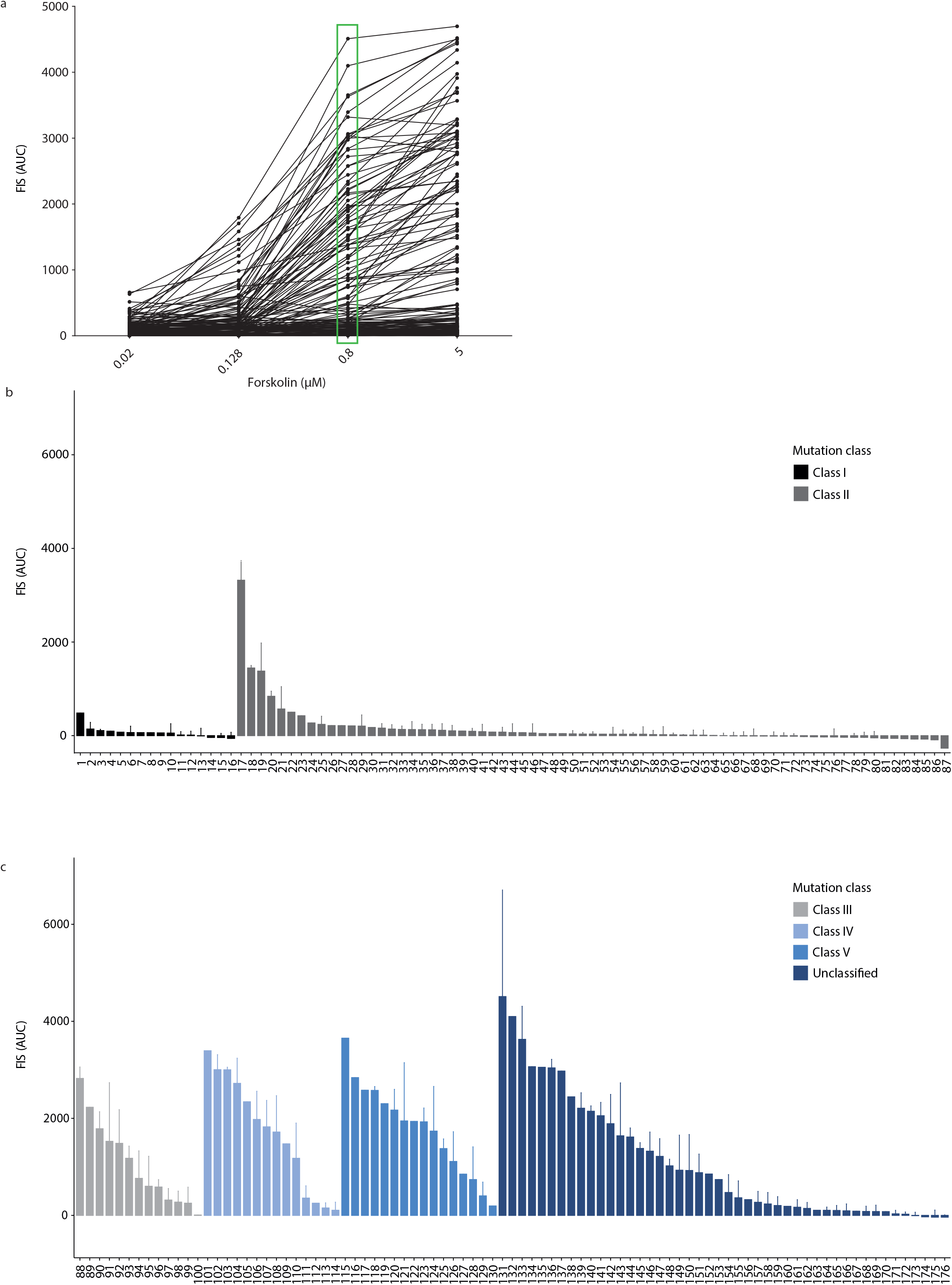
Forksolin induced swelling (FIS) levels of organoids derived from the 176 study participants. a) FIS levels, defined by relative size increase of intestinal organoids after 1h stimulation with four ascending forskolin (fsk) concentrations, quantified as area under the curve (AUC). Each line represent swelling of organoids of individual study participants. Each data point (black dot) represents mean AUC of both technical (n=2) and biological replicates (ranging from n=1-n=7). (b) Waterfall plots of FIS responses at 0.8 µM fsk (highlighted in 1a by the green box) of all study participants grouped based on mutation class I or II or (c) mutation class III-V or unclasified. Genotypes are classified in one mutation class based on the mildest mutation class of the two alleles. Bars represent mean+SD of all replicates. The numbers on the x-axes represent the participant number, the corresponding genotypes are specified in supplemental table 2.

In total, 1072 observations of 152 participants with available FEV1pp measurements (figure 2a) were included in the analysis to assess the association of FIS with long-term FEV1pp decline. Linear mixed model analysis showed that average FEV1pp decline per year of age varied with FIS level, adjusted for sex, genotype class, CFTR modulator usage and SCC (table 1). To illustrate this linear association of FEV1pp decline by age with FIS, figure 2b shows that average annual FEV1pp decline was 1.19 (1.46-0.92, p<0.001) per year of age for participants with a FIS level of 0. Per 1000-points increase in AUC, FEV1pp decline was 0.40 (0.20-0.60, p<0.001) per year of age lower, leading to an FEV1pp decline of 0 for participants with an AUC of 3000.

**Table 1.** Association of FIS with FEV1pp decline. Regression coefficients of linear mixed effects model for FEV1pp.

| N=152, obs=1072 | Coefficient (95% CI) | P-value |
| --- | --- | --- |
| Age | -1.19 (-1.46 - -0.92) | <0.001* |
| FIS | -4.23 (-9.41 – 0.94) | 0.109 |
| FIS*age | 0.40 (0.20 – 0.60) | <0.001* |
| Treatment |  |  |
| - none | Reference category |  |
| - ivacaftor | 7.92 (4.54 – 11.30) | <0.001* |
| - lumacaftor/ivacaftor | -3.87 (-8.29 – 0.55) | 0.086 |
| Sex |  |  |
| - male | Reference category |  |
| - female | -0.77 (-6.70 – 5.15) | 0.798 |
| Genotype class |  |  |
| - unclassified | Reference category |  |
| - class I | -7.43 (-19.26 – 4.40) | 0.218 |
| - class II | -4.71 (-12.69 – 3.27) | 0.247 |
| - class III | -0.46 (-12.26 – 11.35) | 0.939 |
| - class IV | 0.67 (-12.29 – 13.64) | 0.919 |
| - class V | -20.43 (-33.60 – -7.26) | 0.002* |
| SCC | -0.08 (-0.23 – 0.07) | 0.286 |

*FEV1pp: Forced expiratory volume in 1 second, percent predicted. Age in years. FIS: Forskolin induced swelling, defined* *as the relative size increase of intestinal organoids (AUC) after 1h stimulation with 0.8 µM forskolin, AUC scaled 1:1000.*
*FIS\*age indicates the difference in annual FEV1pp decline per 1000 AUC change in FIS level. Genotype class: CFTR-* *protein function class of the mildest of both CFTR-mutations. SCC: Sweat chloride concentration in mmol/L.*
*\* significance level $P < 0.05$ .*

**Figure 2:**
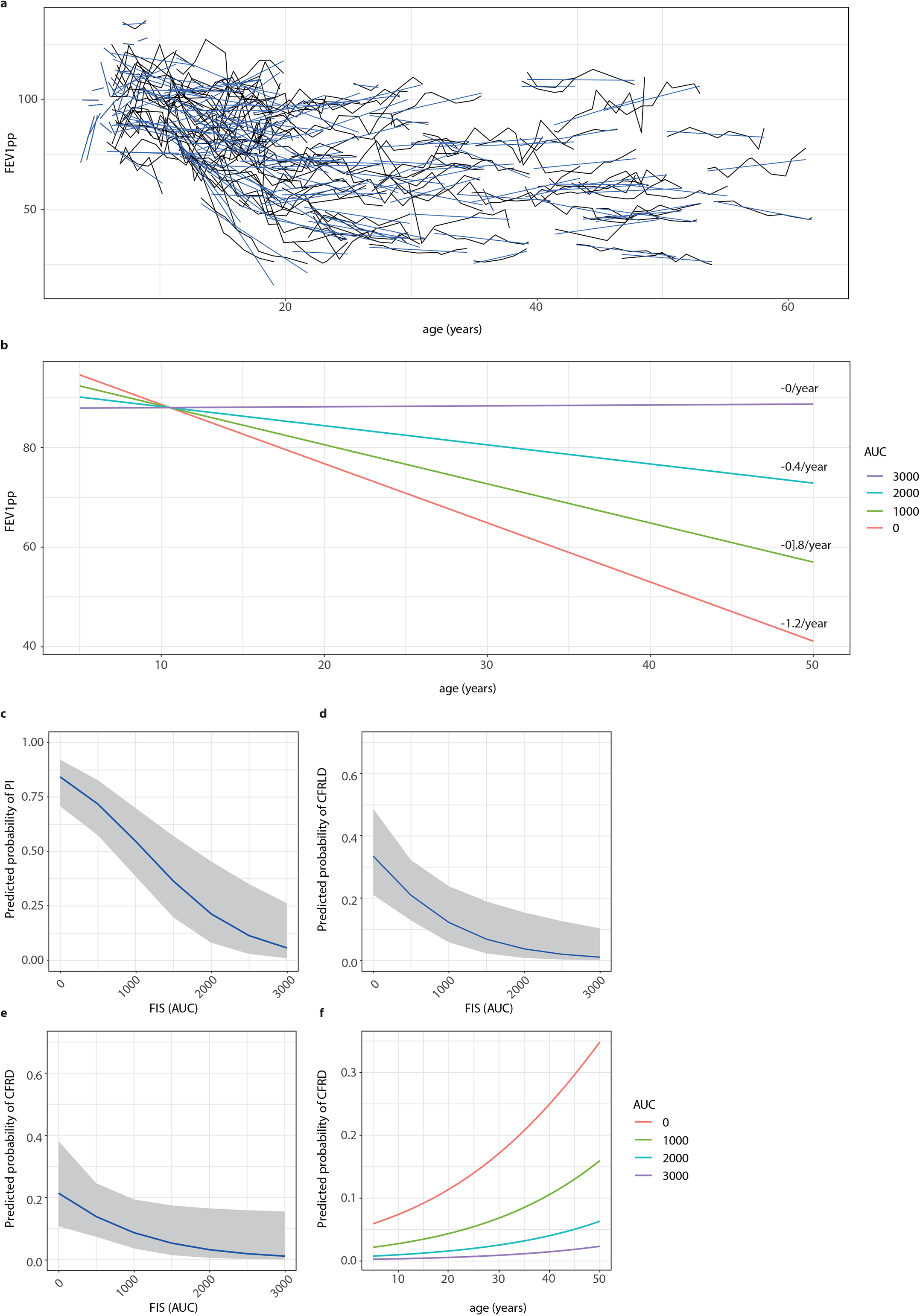
Association of FIS with clinical outcomes. a) individual FEV1pp trajectories of study participants over time in years. b) Association between increasing levels of residual CFTR function (defined by steps of 1000 AUC) and long-term FEV1pp decline for age ranging from 0-50 with the average predicted annual FEV1pp decline specified on the right. (c-e) Association between residual CFTR function (defined by steps of 1000 AUC) and probability of developing pancreas insufficiency (PI) (c), CF-related liver disease (CFRLD) (d) and CF-related diabetes (CFRD) (e). in addition to FIS, age is also associated with probability of developing CFRD (f).

The validity of these results was verified by assessing the potential impact of selection bias and confounding with separate subgroup and sensitivity analyses. A subgroup analysis that assumed a linear relation between age and FEV1pp decline of pwCF between 5-25 years of age identified a similar regression coefficient of 0.49 (0.02-0.96, p=0.039) compared to the complete population, suggesting a negligible impact of selection bias due to inclusion of pwCF with a milder phenotype who survive to an older age. Since at least one *CFTR* mutation was unclassified in 26% of participants (figure 1c, supplementary table 1 and 2), a sensitivity analysis was performed in which we refitted both models with genotype group instead of genotype class, to assess whether the association of FIS with FEV1pp decline was influenced by categorization of genotype. The association of FIS with FEV1pp decline in these models was still statistically significant and coefficients were comparable to the models categorizing genotype by mutation class (supplementary table 4).

In addition, we compared the predictive capacity of FIS with SCC on FEV1pp decline in similar linear mixed models. SCC alone was not significantly associated with FEV1pp decline (p=0.064), although a trend was suggested (supplementary table 5). SCC did not improve the prediction of FEV1pp decline in combination with FIS (p=0.946; supplementary table 6), indicating that FIS predicts long-term FEV1pp decline more accurately than SCC.

To investigate the association of FIS with the occurrence of other CF-related co-morbidities, we performed multivariable logistic regression with PI, CFRLD and CFRD, adjusted for age, sex and SCC. We found a significant association of FIS with the occurrence of PI (adjusted OR: 0.23, 95% CI 0.11-0.49, p<0.001), CFRLD (adjusted OR: 0.28, 95% CI 0.12-0.68, p=0.005) and CFRD (adjusted OR: 0.36, 95% CI 0.13-0.98, p=0.045; table 2 and figure 2c-e). This indicates that the probability of developing CF-related co-morbidities is lower for participants with higher FIS levels. As illustrated in table 2 and figure 2f, age was also significantly associated with the occurrence of CFRD.

**Table 2.** Association of FIS with CF-related co-morbidities. Adjusted odds ratios of multivariable logistic regression for pancreatic insufficiency, CF-related diabetes and CF-related liver disease.

| N=173 | Pancreatic insufficiency |  | CF-related liver disease |  | CF-related diabetes |  |
| --- | --- | --- | --- | --- | --- | --- |
|  | Odds ratio (95% CI) | P-value | Odds ratio (95% CI) | P-value | Odds ratio (95% CI) | P-value |
| FIS | 0.23 (0.11 – 0.49) | <0.001* | 0.28 (0.12 – 0.68) | 0.005* | 0.36 (0.13 – 0.98) | 0.045* |
| Age | 1.00 (0.96 – 1.03) | 0.813 | 1.02 (0.99 – 1.04) | 0.235 | 1.05 (1.02 – 1.08) | 0.003* |
| Sex |  |  |  |  |  |  |
| - male | Reference category |  |  |  |  |  |
| - female | 0.50 (0.18 – 1.36) | 0.184 | 0.72 (0.35 – 1.51) | 0.388 | 2.06 (0.80 – 5.31) | 0.133 |
| SCC | 1.00 (0.97 – 1.02) | 0.754 | 1.01 (0.98 – 1.03) | 0.636 | 1.01 (0.98 – 1.04) | 0.540 |

*FIS: Forskolin induced swelling, defined as the relative size increase of intestinal organoids (AUC) after 1h stimulation* *with 0.8 µM forskolin, AUC scaled 1:1000. Age in years. SCC: sweat chloride concentration in mmol/L.*
*\* significance level $P < 0.05$ .*

In combination with FIS, SCC was clearly not associated with any of the CF-related co-morbidities, given the non-significant odds ratios around 1 (table 2). This suggests that FIS is a stronger predictor of CF-related co-morbidities compared to SCC.

This study showed that residual CFTR function quantified by FIS of patient-derived cystic fibrosis organoids predicts long-term annual FEV1pp decline and occurrence of CF-related co-morbidities PI, CFRLD and CFRD, using 9-year longitudinal data of Dutch pwCF with many distinct *CFTR* mutations and ages ranging from 0 to 61 years old. Despite the influence of genetic modifiers and other non-CFTR dependent environmental factors on CF disease severity ^1,16–18^, it was remarkable to observe that in vitro FIS on intestinal cells has such a broad predictive capacity for many non-intestinal organ systems. Furthermore, direct comparison of FIS with SCC revealed that FIS also predicts multi-organ disease expression better than SCC, which has been the most important and well-validated biomarker of CF disease until now and is a commonly used endpoint to measure efficacy of CFTR modulating drugs ^6,7^. This difference between FIS and SCC could be explained by a more precise and accurate estimation of CFTR function by FIS, since FIS is completely CFTR dependent with minimal impact of technical variability ^11,12^, whereas a substantial part of variability in SCC is caused by technical and other non-CFTR dependent biological factors ^6^. In addition, FIS not only demonstrated a large variability in CFTR function between pwCF with different genotypes but also within genotype classes, illustrating that FIS has the potential to refine the CFTR protein function-based classification system which currently only distinguishes between mild and severe CF phenotypes ^2,19,20^.

Rates of annual FEV1pp decline in this study were within the same range as reported by other recent European studies, which also showed that annual FEV1pp decline was lower for pwCF with a milder disease severity as classified by genotype ^21^ or pancreatic status ^22^ and was highest in the age group between 18 and 28 years ^22^. Moreover, our results are consistent with a previous study showing a more severe CF disease phenotype in terms of pulmonary and gastrointestinal outcome parameters in infants with low FIS compared to infants with high FIS ^13^. In line with our observations, Davis et al. also demonstrated that SCC by itself does not predict lung disease in pwCF ^23^.

An important limitation of this research is the retrospective observational study design. We adjusted for several confounders, but were unable to account for other prognostic factors such as pulmonary exacerbations and sputum cultures. As 34% of SCC values was missing, we used multiple imputation methods to prevent bias due to selective missing data. Potential impact of survival bias was minimized by our subgroup and sensitivity analyses, but could not completely be excluded. Further research is also needed to study the association of changes in organoid FIS with long-term clinical effects upon CFTR modulator therapy.

In summary, this study showed that FIS of cystic fibrosis organoids is strongly associated with long-term FEV1pp decline and occurrence of different CF-related co-morbidities, suggesting that estimation of CFTR function by FIS could have important prognostic value for individual disease expression in multiple organs that affect CF morbidity.

## Data Availability

The datasets generated and analyzed in the current study are available from the corresponding author on reasonable request.

## Acknowledgements

This work was supported by grants of the Dutch Cystic Fibrosis Foundation (NCFS) as part of the HIT-CF Program and by ZonMW. Furthermore, we would like to thank the people with CF who gave informed consent for generating and testing their individual organoids and the use of their data; all members of the research teams of the Dutch CF clinics that contributed to this work; and all colleagues of the HUB Organoid Technology for their help with generating intestinal organoids and performing FIS experiments.

## Author Contribution Statement

D.M. and E.d.P. contributed substantially to the design of the study, the acquisition, analysis and interpretation of data and have drafted the manuscript.

S.W.F.S., A.M.V., J.E.B., E.K., H.O., M.C.H., P.v.M. G.B., K.M.d.W-d.G., S.H.-M., S.R.J., H.v.P., M.M.v.d.E., R.v.d.M., J.R., E.D., E.J.M.W., G.H.K., R.V. and D.D.Z.-v.O. contributed to the acquisition of study data and revised the manuscript. M.J.C.E. contributed to the design of the study, analysis and interpretation of data and revised the manuscript. C.K.v.d.E and J.M.B. have made substantial contributions to the conception and design of the study, interpretation of data and revised the manuscript.

## Conflict of interest

J.M.B. and C.K.v.d.E. are inventors on patent(s) related to the FIS-assay and received financial royalties from 2017 onward. J.M.B report receiving research grant(s) and consultancy fees from various industries, including Vertex Pharmaceuticals, Proteostasis Therapeutics, Eloxx Pharmaceuticals, Teva Pharmaceutical Industries and Galapagos outside the submitted work. C.K.v.d.E report receiving research grant(s) grant(s) from Vertex Pharmaceuticals (money to institution) outside the submitted work. G.H.K. reports grants from Lung Foundation of the Netherlands, Vertex Pharmaceuticals, UBBO EMMIUS foundation, GSK, TEVA the Netherlands, TETRI Foundation, European Union (H2020) (all money to institution), outside the submitted work.

## Code availability statement

No custom code or mathematical algorithm was used for this study.

## Material availability statement

Intestinal organoids are accessible for study by contacting the Hubrecht Organoid Technology foundation (http://hub4organoids.eu/). Further information and requests for resources and reagents should be directed to and will be fulfilled by the lead contact, Dr. Jeffrey M. Beekman.

## Clinical data

Clinical data was retrieved from the Dutch Cystic Fibrosis Foundation (NCFS) of participants who provided written informed consent.

## MATERIALS & METHODS

### Study design and population

A retrospective observational cohort study was conducted in Dutch people with CF (pwCF) of which 1) intestinal organoids were generated as part of previous studies or clinical care before January 2020 and 2) written informed consent was obtained to use their intestinal organoids and clinical data for the present study.

### Study parameters

The primary outcome variable was defined as long-term lung function decline, expressed as FEV1 percent predicted (FEV1pp) calculated according to global lung function initiative (GLI) guidelines. Secondary outcome variables were occurrence of pancreatic insufficiency (PI), defined by fecal elastase < 200 µg/g, clinical diagnosis of CF-related liver disease (CFRLD) and occurrence of insulin-dependent CF-related diabetes (CFRD) defined by daily insulin treatment.

The primary explanatory variable of interest was forskolin induced swelling (FIS), defined by the relative size increase of intestinal organoids after 1h stimulation with 0.8 µM forskolin (fsk), quantified as area under the curve (AUC). Previous studies showed that discrimination between individual FIS responses was most optimal and correlated best with other in vitro and in vivo CFTR biomarkers when FIS was performed with 0.8 µM fsk ^12,13^. Other included explanatory variables were: age in years at time of each lung function measurement; treatment status at time of each lung function measurement, categorized as no CFTR-modulator treatment, treatment with ivacaftor or with lumacaftor/ivacaftor; sex; sweat chloride concentration (SCC) in mmol/L; and genotype, categorized as class I-V or unclassified, defined by genotype class of the mildest of both mutations according to available literature (supplementary table 2). Additionally, genotypes were categorized in groups according to the combination of the following mutation types: insertion/deletion, nonsense, missense, splice, unknown.

### Study procedures

#### Organoid measurements

The generation of intestinal organoids from biopsies and subsequent fluid secretion assays (FIS-assays) were performed according to a previously described protocol ^34^. All FIS-assay experiments were conducted in duplicate and for the majority of the donors at multiple culturing time points with a maximum of 7 consecutive culture time points (n=7).

#### Clinical data collection

Data on clinical study parameters were retrieved from the Dutch Cystic Fibrosis Foundation (NCFS) Registry. Annual best FEV1pp values between 2010 and 2018 were used to estimate lung function decline. Treatment status at time of lung function measurements was calculated based on start and stop dates of CFTR-modulators registered in the database. Of all other variables, single values known in 2018 were obtained because repeated measurements were unavailable or inconsistently collected.

### Statistical analysis

Multilevel multiple imputation was performed to handle missing data of SCC, PI and CFRD under the assumption of missingness at random (MAR). All statistical analyses were performed on multiple imputed datasets (m=5, iterations=20) with pooling of the results.

The association between age and long-term lung function decline was analyzed using a linear mixed effects model. FEV1pp was specified as outcome variable in the model, with FIS, SCC, genotype class, sex, age, treatment and FIS*age as fixed effects, where the interaction term FIS*age reflected the difference in annual FEV1pp decline according to FIS level. The model included a random intercept and random slope for age per subject, assuming a first order auto-regressive (cAR1) correlation structure.

To account for selection bias towards a milder phenotype in pwCF surviving to an older age, a subgroup analysis was conducted only including measurements between 5-25 years, in which the relationship between age and FEV1pp decline can reasonably be assumed to be linear.

Sensitivity analyses were performed using genotype group instead of genotype class, to assess whether the association of FIS with FEV1pp decline was influenced by categorization of genotype. To obtain reliable effect estimates and standard errors for genotype group, groups with less than 5 participants were excluded from this part of the analysis.

To compare the predictive value of FIS with SCC on long-term FEV1pp decline, four models were built which all included FIS, SCC, genotype class, sex, age and treatment as fixed effects. A baseline model was built without an interaction term, and the other three models were built with the addition of either the interaction term FIS*age, SCC*age or both FIS*age and SCC*age in the model. Performance of these models was compared using the Likelihood Ratio test.

Secondary outcomes were analyzed using multivariable logistic regression, with FIS, SCC, sex, and age in 2018 as explanatory variables. Given the low proportion of outcome events within some of the genotype classes as well as within genotype groups, genotype was not included in the analysis. In addition, CFTR modulator usage was not included because date of onset of the secondary outcomes was not available, indicating that a causal effect could not be addressed.

Significance levels were set at 0.05. Statistical analyses were performed with R version 4.0.2 using packages nlme, lme4, mice and micemd.

## Notes

### Author Declarations

Storage and use of intestinal organoids and the collection and linking of clinical data was approved by the Institutional Review Board (IRB) of the University Medical Center Utrecht (UMCU), The Netherlands and registered under number 14-008 (TC Biobank) and 16-668C (CCFR cohort).

## REFERENCES

1. Cutting, G. R. Cystic fibrosis genetics: From molecular understanding to clinical application. Nat. Rev. Genet. 16, 45–56 (2015).

2. US CF Foundation, Johns Hopkins University, T. H. for S. C. The Clinical and Functional TRanslation of CFTR (CFTR2). http://cftr2.org (2011).

3. Elborn, J. S. Cystic fibrosis. Lancet (London, England) 388, 2519–2531 (2016).

4. Beekman, J. M. et al. CFTR functional measurements in human models for diagnosis, prognosis and personalized therapy: Report on the pre-conference meeting to the 11th ECFS Basic Science Conference, Malta, 26-29 March 2014. J. Cyst. Fibros. Off. J. Eur. Cyst. Fibros. Soc. 13, 363–372 (2014).

5. Bronsveld, I. et al. Residual chloride secretion in intestinal tissue of deltaF508 homozygous twins and siblings with cystic fibrosis. The European CF Twin and Sibling Study Consortium. Gastroenterology 119, 32–40 (2000).

6. Collaco, J. M. et al. Sources of Variation in Sweat Chloride Measurements in Cystic Fibrosis. Am. J. Respir. Crit. Care Med. 194, 1375–1382 (2016).

7. De Boeck, K. et al. CFTR biomarkers: time for promotion to surrogate end-point. Eur. Respir. J. 41, 203–216 (2013).

8. Kyrilli, S. et al. Insights into the variability of nasal potential difference, a biomarker of CFTR activity. J. Cyst. Fibros. Off. J. Eur. Cyst. Fibros. Soc. 19, 620–626 (2020).

9. Mall, M. et al. The DeltaF508 mutation results in loss of CFTR function and mature protein in native human colon. Gastroenterology 126, 32–41 (2004).

10. Minso, R. et al. Intestinal current measurement and nasal potential difference to make a diagnosis of cases with inconclusive CFTR genetics and sweat test. BMJ open Respir. Res. 7, (2020).

11. Dekkers, J. F. et al. A functional CFTR assay using primary cystic fibrosis intestinal organoids. Nat. Med. 19, 939–945 (2013).

12. Dekkers, J. F. et al. Characterizing responses to CFTR-modulating drugs using rectal organoids derived from subjects with cystic fibrosis. Sci. Transl. Med. 8, 344ra84 (2016).

13. de Winter-de Groot, K. M. et al. Stratifying infants with cystic fibrosis for disease severity using intestinal organoid swelling as a biomarker of CFTR function. Eur. Respir. J. 52, (2018).

14. de Winter-de Groot, K. M. et al. Forskolin-induced swelling of intestinal organoids correlates with disease severity in adults with cystic fibrosis and homozygous F508del mutations. J. Cyst. Fibros. Off. J. Eur. Cyst. Fibros. Soc. 19, 614–619 (2020).

15. Ramalho, A. S. et al. Correction of CFTR function in intestinal organoids to guide treatment of cystic fibrosis. Eur. Respir. J. 57, (2021).

16. Collaco, J. M., Blackman, S. M., McGready, J., Naughton, K. M. & Cutting, G. R. Quantification of the relative contribution of environmental and genetic factors to variation in cystic fibrosis lung function. J. Pediatr. 157, 802–803 (2010).

17. Santis, G., Osborne, L., Knight, R. A. & Hodson, M. E. Independent genetic determinants of pancreatic and pulmonary status in cystic fibrosis. Lancet (London, England) 336, 1081–1084 (1990).

18. Weiler, C. A. & Drumm, M. L. Genetic influences on cystic fibrosis lung disease severity. Front. Pharmacol. 4, 40 (2013).

19. McKone, E. F., Emerson, S. S., Edwards, K. L. & Aitken, M. L. Effect of genotype on phenotype and mortality in cystic fibrosis: a retrospective cohort study. Lancet (London, England) 361, 1671–1676 (2003).

20. McKone, E. F., Goss, C. H. & Aitken, M. L. CFTR genotype as a predictor of prognosis in cystic fibrosis. Chest 130, 1441–1447 (2006).

21. De Boeck, K. & Zolin, A. Year to year change in FEV(1) in patients with cystic fibrosis and different mutation classes. J. Cyst. Fibros. Off. J. Eur. Cyst. Fibros. Soc. 16, 239–245 (2017).

22. Caley, L., Smith, L., White, H. & Peckham, D. G. Average rate of lung function decline in adults with cystic fibrosis in the United Kingdom: Data from the UK CF registry. J. Cyst. Fibros. Off. J. Eur. Cyst. Fibros. Soc. (2020) doi:10.1016/j.jcf.2020.04.008.

23. Davis, P. B., Schluchter, M. D. & Konstan, M. W. Relation of sweat chloride concentration to severity of lung disease in cystic fibrosis. Pediatr. Pulmonol. 38, 204–209 (2004).

24. De Boeck, K., Zolin, A., Cuppens, H., Olesen, H. V & Viviani, L. The relative frequency of CFTR mutation classes in European patients with cystic fibrosis. J. Cyst. Fibros. Off. J. Eur. Cyst. Fibros. Soc. 13, 403–409 (2014).

25. Green, D. M. et al. Mutations that permit residual CFTR function delay acquisition of multiple respiratory pathogens in CF patients. Respir. Res. 11, 140 (2010).

26. Yu, H. et al. Ivacaftor potentiation of multiple CFTR channels with gating mutations. J. Cyst. Fibros. Off. J. Eur. Cyst. Fibros. Soc. 11, 237–245 (2012).

27. Vankeerberghen, A. et al. Characterization of mutations located in exon 18 of the CFTR gene. FEBS Lett. 437, 1–4 (1998).

28. Ren, H. Y. et al. VX-809 corrects folding defects in cystic fibrosis transmembrane conductance regulator protein through action on membrane-spanning domain 1. Mol. Biol. Cell 24, 3016–3024 (2013).

29. Rapino, D. et al. Rescue of NBD2 mutants N1303K and S1235R of CFTR by small-molecule correctors and transcomplementation. PLoS One 10, e0119796 (2015).

30. Pagani, F. et al. New type of disease causing mutations: the example of the composite exonic regulatory elements of splicing in CFTR exon 12. Hum. Mol. Genet. 12, 1111–1120 (2003).

31. Van Goor, F., Yu, H., Burton, B. & Hoffman, B. J. Effect of ivacaftor on CFTR forms with missense mutations associated with defects in protein processing or function. J. Cyst. Fibros. Off. J. Eur. Cyst. Fibros. Soc. 13, 29–36 (2014).

32. De Boeck, K. Cystic fibrosis in the year 2020: A disease with a new face. Acta Paediatr. 109, 893–899 (2020).

33. Prontera, P. et al. A Clinical and Molecular Survey of 62 Cystic Fibrosis Patients from Umbria (Central Italy) Disclosing a High Frequency (2.4%) of the 2184insA Allele: Implications for Screening. Public Health Genomics 19, 336–341 (2016).

34. Vonk, A. M. et al. Protocol for Application, Standardization and Validation of the Forskolin-Induced Swelling Assay in Cystic Fibrosis Human Colon Organoids. STAR Protoc. 1, 100019 (2020).

