## Supplementary figure and tables for "Forskolin-induced swelling of intestinal organoids predicts long-term cystic fibrosis disease progression"

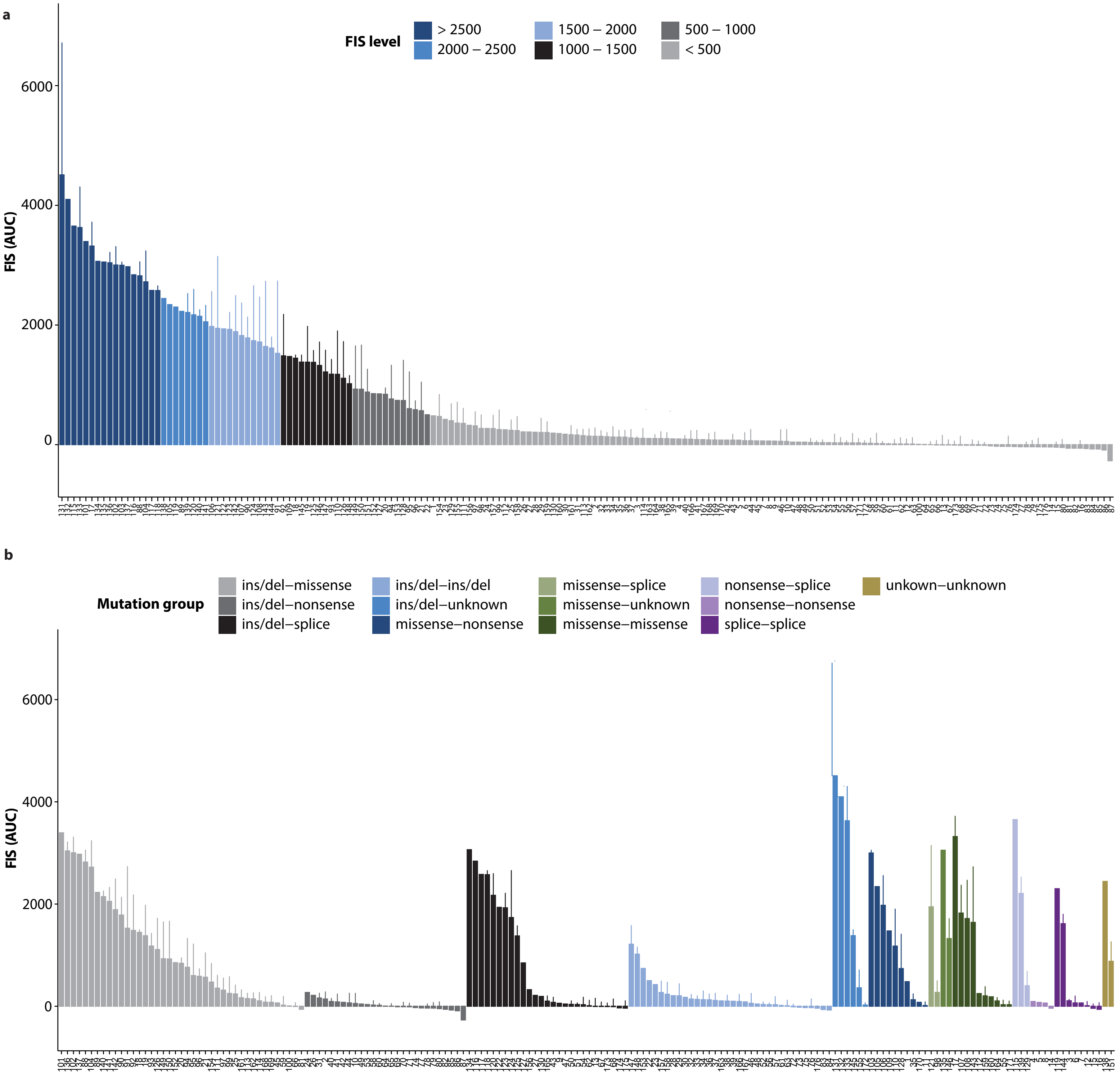


**Supplementary Figure 1**: a) waterfall plot of FIS responses stimulated with 0.8 µM fsk for one hour of all study participants. b) waterfall plot of FIS responses at 0.8 µM fsk per mutation group. Groups were defined by the combination of the mutation type of both mutations. Bars represent mean+SD of replicates, ranging from n=2 to n=7. The numbers on the x-axes represent the participant number and correspond to the numbers in figure 1. Genotypes are specified in supplementary table 2.

**Supplementary information:**

**Supplementary table 1. Participant characteristics**

| **N=173** | |
|---|---|
| **Age, median (IQR)** | 20.2 (9.4-31.8) |
| **Sex, n (%)** | |
| Male | 89 (51) |
| Female | 84 (49) |
| **Mutation class, n (%)** | |
| Class I | 16 (9.2) |
| Class II | 71 (41.0) |
| Class III | 13 (7.5) |
| Class IV | 14 (8.1) |
| Class V | 16 (9.2) |
| Unclassified | 43 (24.9) |
| **CFTR-modulator use, n (%)** | |
| Ivacaftor | 16 (9.2) |
| Lumacaftor/ivacaftor | 8 (4.6) |
| **FIS, median (IQR)** | 142.5 (30.3-1214.9) |
| **SCC, mean (SD)** | 92.2 (26.2) |
| Missing values, n (%) | 58 (34) |
| **FEV1pp, mean (SD)** | 79.1 (24.9) |
| **Pancreatic function, n (%)** | |
| Insufficient (fecal elastase <200 µg/g) | 77 (45) |
| Sufficient (fecal elastase ≥200 µg/g) | 35 (20) |
| Missing values | 61 (35) |
| **CF-related liver disease, n (%)** | 45 (26) |
| Missing values | 2 (1) |
| **CF-related diabetes (insulin-dependent), n (%)** | 25 (14) |

*Age in years. Genotype: genotype class of the mildest of both mutations. FIS: Forskolin induced swelling, defined as the relative size increase of intestinal organoids (AUC) after 1h stimulation with 0.8 µmol/L forskolin, AUC scaled 1:1000. SCC: Sweat chloride concentration in mmol/L. FEV1pp: Forced expiratory volume in 1 second, percent predicted in 2018.*

**Supplementary table 2. Individual genotypes of study participants**

Overview of the individual genotypes with corresponding *CFTR* mutation classification according to available literature. Study participants were categorized into one mutation class based on the mildest of both mutations or to unclassified when one of the mutation classes was unknown. The mutation group was defined by the combination of mutation types of both alleles.

| ID | Genotype | Classification | Mutation group | ID | Genotype | Classification | Mutation group |
|---|---|---|---|---|---|---|---|
| **1** | G542X/R1066C | **I** [24]/ I [25] | Nonsense-missense | **89** | F508del/R75Q | II [24]/ **III** [25] | Ins/del-missense |
| **2** | G542X/CFTRdele2.3 | **I** [24]/ I [24] | Nonsense-ins/del | **90** | F508del/S1251N | II [24]/ **III** [24] | Ins/del-missense |
| **3** | 1811+1G>C/1811+1G>C | **I** [24]/ I [24] | Splice-splice | **91** | F508del/S1251N | II [24]/ **III** [24] | Ins/del-missense |
| **4** | W1282X/W1282X | **I** [24]/ I [24] | Nonsense-nonsense | **92** | F508del/S1251N | II [24]/ **III** [24] | Ins/del-missense |
| **5** | G542X/W679X | **I** [24]/ I [24] | Nonsense-nonsense | **93** | F508del/S1251N | II [24]/ **III** [24] | Ins/del-missense |
| **6** | 1811+1G>C/1811+1G>C | **I** [24]/ I [24] | Splice-splice | **94** | F508del/S1251N | II [24]/ **III** [24] | Ins/del-missense |
| **7** | 3849+10kbC>T/1717-1G>A | **I** [24]/ I [24] | Splice-splice | **95** | F508del/S1251N | II [24]/ **III** [24] | Ins/del-missense |
| **8** | R785X/R785X | **I** [24]/ I [24] | Nonsense-nonsense | **96** | F508del/S1251N | II [24]/ **III** [24] | Ins/del-missense |
| **9** | 1717-1G>A/3905insT | **I** [24]/ I [25] | Splice-ins/del | **97** | F508del/S1251N | II [24]/ **III** [24] | Ins/del-missense |
| **10** | R1162X/3659delC | **I** [24]/ I [24] | Nonsense-ins/del | **98** | S1251N/1717-1G>A | **III** [24]/ I [24] | Ins/del-splice |
| **11** | G542X/R1066C | **I** [24]/ I [25] | Nonsense-missense | **99** | F508del/S1251N | II [24]/ **III** [24] | Ins/del-missense |
| **12** | 1811+1G>C/1811+1G>C | **I** [24]/ I [24] | Splice-splice | **100** | F508del/G178R | II [24]/ **III** [26] | Ins/del-missense |
| **13** | 711+1G>T/CFTRdele11 | **I** [24]/ I [24] | Splice-ins/del | **101** | 3905insT/D1152H | I [25]/ **IV** [27] | Ins/del-missense |
| **14** | L732X/L732X | **I** [24]/ I [24] | Nonsense-nonsense | **102** | F508del/D1152H | II [24]/ **IV** [27] | Ins/del-missense |
| **15** | 1811+1G>C/1811+1G>C | **I** [24]/ I [24] | Splice-splice | **103** | W1282X/R117H;7T | I [24]/ **IV** [24] | Nonsense/missense |
| **16** | 711+1G>T/711+1G>T | **I** [24]/ I [24] | Splice-splice | **104** | F508del/R117H;7T/9T | II [24]/ **IV** [24] | Ins/del-missense |
| **17** | L206W/S1235R | **II** [28]/ II [29] | Missense-missense | **105** | R1162X/D1152H | I [24]/ **IV** [27] | Nonsense/missense |
| **18** | F508del/G628R | **II** [24]/ II [30] | Ins/del-missense | **106** | R117H;7T/R553X | **IV** [24]/ I [24] | Missense/nonsense |
| **19** | F508del/I336K | **II** [24]/ II [31] | Ins/del-missense | **107** | R334W/N1303K | **IV** [24]/ II [24] | Missense-missense |
| **20** | F508del/G628R | **II** [24]/ II [24] | Ins/del-missense | **108** | R334W/R334W | **IV** [24]/ IV [24] | Missense-missense |
| **21** | F508del/G628R | **II** [24]/ II [24] | Ins/del-missense | **109** | D1152H/R1162X | **IV** [24]/ I [27] | Missense-nonsense |
| **22** | F508del/F508del | **II** [24]/ II [24] | Ins/del-ins/del | **110** | R334W/R764X | **IV** [24]/ I [24] | Missense-nonsense |

| | | | | | | | |
|---|---|---|---|---|---|---|---|
| **23** | F508del/2184delA | **II** [24]/ I [25] | Ins/del-ins/del | **111** | F508del/R347P | II [24]/ **IV** [24] | Ins/del-missense |
| **24** | F508del/Y1092X | **II** [24]/ I [24] | Ins/del-nonsense | **112** | L1335P/L1335P | **IV** [25]/ IV [25] | Missense-missense |
| **25** | F508del/R1066C | **II** [24]/ I [25] | Ins/del-missense | **113** | F508del/R347P | II [24]/ **IV** [24] | Ins/del-missense |
| **26** | F508del/W846X | **II** [24]/ I [24] | Ins/del-nonsense | **114** | F508del/R347P | II [24]/ **IV** [24] | Ins/del-missense |
| **27** | F508del/1717-1G>A | **II** [24]/ I [24] | Ins/del-splice | **115** | G542X/3849+10kbC>T | I [24]/ **V** [24] | Nonsense-splice |
| **28** | F508del/CFTRdele17a.17b | **II** [24]/ I [24] | Ins/del-ins/del | **116** | F508del/3849+10kbC>T | II [24]/ **V** [24] | Ins/del-splice |
| **29** | F508del/F508del | **II** [24]/ II [24] | Ins/del-ins/del | **117** | F508del/3849+10kbC>T | II [24]/ **V** [24] | Ins/del-splice |
| **30** | F508del/1078delT | **II** [24]/ I [25] | Ins/del-ins/del | **118** | F508del/3849+10kbC>T | II [24]/ **V** [24] | Ins/del-splice |
| **31** | F508del/W1282X | **II** [24]/ I [24] | Ins/del-nonsense | **119** | 3272-26A>G/3272-26A>G | **V** [32]/ V [24] | Splice-splice |
| **32** | F508del/1078delT | **II** [24]/ I [25] | Ins/del-ins/del | **120** | F508del/3849+10kbC>T | II [24]/ **V** [24] | Ins/del-splice |
| **33** | F508del/F508del | **II** [24]/ II [24] | Ins/del-ins/del | **121** | 3272-26A>G/G970R | **V** [32]/ III [26] | Splice-missense |
| **34** | F508del/F508del | **II** [24]/ II [24] | Ins/del-ins/del | **122** | F508del/3272-26A>G | II [24]/ **V** [32] | Ins/del-splice |
| **35** | N1303K/G550X | **II** [24]/ I [24] | Missense-nonsense | **123** | F508del/3272-26A>G | II [24]/ **V** [32] | Ins/del-splice |
| **36** | F508del/CFTRdele19.20 | **II** [24]/ I [24] | Ins/del-ins/del | **124** | F508del/3272-26A>G | II [24]/ **V** [32] | Ins/del-splice |
| **37** | F508del/F508del | **II** [24]/ II [24] | Ins/del-ins/del | **125** | F508del/3272-26A>G | II [24]/ **V** [32] | Ins/del-splice |
| **38** | F508del/3659delC | **II** [24]/ I [24] | Ins/del-ins/del | **126** | A455E/3659delC | **V** [24]/ I [24] | Missense-ins/del |
| **39** | F508del/CFTRdele17a.17b | **II** [24]/ I [24] | Ins/del-ins/del | **127** | F508del/3272-26A>G | II [24]/ **V** [32] | Ins/del-splice |
| **40** | F508del/E730X | **II** [24]/ I [24] | Ins/del-nonsense | **128** | A455E/E60X | **V** [24]/ I [24] | Missense-nonsense |
| **41** | F508del/Y1092X | **II** [24]/ I [24] | Ins/del-nonsense | **129** | Y849X/2789+5G>A | I [24]/ **V** [24] | Nonsense-splice |
| **42** | F508del/E60X | **II** [24]/ I [24] | Ins/del-nonsense | **130** | 1078delT/3272-26A>G | I [25] / **V** [32] | Ins/del-splice |
| **43** | F508del/711+1G>T | **II** [24]/ I [24] | Ins/del-splice | **131** | F508del/(TG)13(T)5 | II [24]/ **unclassified** | Ins/del-unknown |
| **44** | F508del/Y1092X | **II** [24]/ I [24] | Ins/del-nonsense | **132** | F508del/(TG)13(T)5 | II [24]/ **unclassified** | Ins/del-unknown |
| **45** | F508del/G85E | **II** [24]/ II [24] | Ins/del-missense | **133** | F508del/(TG)13(T)5 | II [24]/ **unclassified** | Ins/del-unknown |
| **46** | F508del/I507del | **II** [24]/ II [24] | Ins/del-ins/del | **134** | F508del/c.4243-3T>A | II [24]/ **unclassified** | Ins/del-splice |
| **47** | F508del/1717-1G>T | **II** [24]/ I [24] | Ins/del-splice | **135** | A455E/(TG)13(T)5 | V [24]/ **unclassified** | Missense-unknown |
| **48** | F508del/3659delC | **II** [24]/ I [24] | Ins/del-ins/del | **136** | F508del/R1358S | II [24]/ **unclassified** | Ins/del-missense |
| **49** | F508del/W1282X | **II** [24]/ I [24] | Ins/del-nonsense | **137** | F508del/G576A | II [24]/ **unclassified** | Ins/del-missense |
| **50** | F508del/711+1G>T | **II** [24]/ I [24] | Ins/del-splice | **138** | Unknown/unknown | **unclassified**/ unclassified | Unknown-unknown |
| **51** | F508del/1717-1G>A | **II** [24]/ I [24] | Ins/del-splice | **139** | R553X/c.4375-3T>A | I [24]/ **unclassified** | Nonsense-splice |

| | | | | | | | |
|---|---|---|---|---|---|---|---|
| **52** | F508del/CFTRdele17a.17b | **II** [24]/ I [24] | Ins/del-ins/del | **140** | F508del/L206W | II [24]/ **unclassified** | Ins/del-missense |
| **53** | F508del/Y849X | **II** [24]/ I [24] | Ins/del-nonsense | **141** | F508del/T1396P | II [24]/ **unclassified** | Ins/del-missense |
| **54** | F508del/1717-1G>A | **II** [24]/ I [24] | Ins/del-splice | **142** | F508del/G461R | II [24]/ **unclassified** | Ins/del-missense |
| **55** | N1303K/G85E | **II** / II [24] | Missense-missense | **143** | N1303K/c.3035A>C | II [24]/ **unclassified** | Missense-missense |
| **56** | F508del/CFTRdele2.3 | **II** [24]/ I [24] | Ins/del-ins/del | **144** | 3272-26A>G/1898+5G>T | V [24]/ **unclassified** | Splice-splice |
| **57** | F508del/CFTRdele17a.17b | **II** [24]/ I [24] | Ins/del-ins/del | **145** | F508del/unknown | II [24]/ **unclassified** | Ins/del-unknown |
| **58** | F508del/Y849X | **II** [24]/ I [24] | Ins/del-nonsense | **146** | R117H;7T/unknown | IV [24]/ **unclassified** | Missense-unknown |
| **59** | F508del/G85E | **II** [24]/ II [24] | Ins/del-missense | **147** | 4382delA/2043delG | **unclassified** / unclassified | Ins/del-ins/del |
| **60** | F508del/S489X | **II** [24]/ I [24] | Ins/del-nonsense | **148** | F508del/4382delA | II [24]/ **unclassified** | Ins/del-ins/del |
| **61** | F508del/2184delA | **II** [24]/ I [25] | Ins/del-ins/del | **149** | F508del/G1249R | II /[24] **unclassified** | Ins/del-missense |
| **62** | F508del/711+1G>T | **II** [24]/ I [24] | Ins/del-splice | **150** | A455E/1343delG | V [24]/ **unclassified** | Missens-ins/del |
| **63** | F508del/F508del | **II** [24]/ II [24] | Ins/del-ins/del | **151** | Unknown/unknown | **unclassified** / unclassified | Unknown-unknown |
| **64** | F508del/W1282X | **II** [24]/ I [24] | Ins/del-nonsense | **152** | R1066H/CFTRdele2.3 | **unclassified** / I [24] | Missense-ins/del |
| **65** | F508del/Y1092X | **II** [24]/ I [24] | Ins/del-nonsense | **153** | F508del/4382delA | II [24]/ **unclassified** | Ins/del-ins/del |
| **66** | F508del/N1303K | **II** [24]/ II [24] | Ins/del-missense | **154** | F508del/G1249R | II [24]/ **unclassified** | Ins/del-missense |
| **67** | F508del/711+1G>T | **II** [24]/ I [24] | Ins/del-splice | **155** | F508del/unknown | II [24]/ **unclassified** | Ins/del-unknown |
| **68** | F508del/711+1G>T | **II** [24]/ I [24] | Ins/del-splice | **156** | F508del/3849+5G>T | II [24]/ **unclassified** | Ins/del-splice |
| **69** | F508del/R1162X | **II** [24]/ I [24] | Ins/del-nonsense | **157** | F508del/365-366insT(W79fs) | II [24]/ **unclassified** | Ins/del-ins/del |
| **70** | F508del/G550X | **II** [24]/ I [24] | Ins/del-nonsense | **158** | F508del/1342-1delG | II [24]/ **unclassified** | Ins/del-ins/del |
| **71** | F508del/G550X | **II** [24]/ I [24] | Ins/del-nonsense | **159** | R1066C/R1066H | II [25]/ **unclassified** | Missense-missense |
| **72** | F508del/F508del | **II** [24]/ II [24] | Ins/del-ins/del | **160** | V1160T/E92K | **unclassified** / II [24] | Missense-missense |
| **73** | F508del/F508del | **II** [24]/ II [24] | Ins/del-ins/del | **161** | F508del/R74P | II [24]/ **unclassified** | Ins/del-missense |
| **74** | F508del/Q493X | **II** [24]/ I [24] | Ins/del-nonsense | **162** | F508del/L1034P | II [24]/ **unclassified** | Ins/del-missense |
| **75** | F508del/4016insT | **II** [24]/ I [25] | Ins/del-ins/del | **163** | F508del/Ile336fs | II [24]/ **unclassified** | Ins/del-missense |
| **76** | F508del/394delTT | **II** [24]/ I [25] | Ins/del-ins/del | **164** | A46D/A46D | **unclassified** / unclassified | Missense-missense |
| **77** | F508del/G550X | **II** [24]/ I [24] | Ins/del-nonsense | **165** | 1717-1G>A/2183AA>G | I [24]/ **unclassified** | Splice-ins/del |
| **78** | F508del/R1162X | **II** [24]/ I [24] | Ins/del-nonsense | **166** | F508del/2183AA>G | II [24]/ **unclassified** | Ins/del-ins/del |
| **79** | F508del/R1162X | **II** [24]/ I [24] | Ins/del-nonsense | **167** | F508del/1813insC | II [24]/ **unclassified** | Ins/del-ins/del |
| **80** | F508del/Y1092X | **II** [24]/ I [24] | Ins/del-nonsense | **168** | F508del/S18I | II [24]/ **unclassified** | Ins/del-missense |

| | | | | | | | |
|---|---|---|---|---|---|---|---|
| **81** | F508del/N1303K | **II** [24]/ II [24] | Ins/del-missense | **169** | F508del/Y109D | II [24]/ **unclassified** | Ins/del-missense |
| **82** | F508del/W1282X | **II** [24]/ I [24] | Ins/del-nonsense | **170** | W1282X/L927P | I [24]/ **unclassified** | Nonsense-missense |
| **83** | F508del/2184insA | **II** [24]/ I [33] | Ins/del-ins/del | **171** | A46D/A46D | **unclassified** / **unclassified** | Missense-missense |
| **84** | F508del/F508del | **II** [24]/ II [24] | Ins/del-ins/del | **172** | F508del/unknown | II [24]/ **unclassified** | Ins/del-unknown |
| **85** | F508del/R1162X | **II** [24]/ I [24] | Ins/del-nonsense | **173** | I507del/c.4242+2T>C | II [24]/ **unclassified** | Ins/del-splice |
| **86** | F508del/E60X | **II** [24]/ I [24] | Ins/del-nonsense | **174** | F508del/IVS11-1G>C | II [24]/ **unclassified** | Ins/del-splice |
| **87** | F508del/R1162X | **II** [24]/ I [24] | Ins/del-nonsense | **175** | 1677delTA/IVS16+1G>A(3120+1G>A) | **unclassified** / I [24] | Ins/del-splice |
| **88** | F508del/I1027T | II [24]/ **III** [25] | Ins/del-missense | **176** | F508del/Gly1349fs | II [24]/ **unclassified** | Ins/del-ins/del |

**Supplementary table 3. Association of FIS with FEV1pp decline in subgroup analysis**

Regression coefficients linear mixed model FEV1pp within a subgroup only including participants with age between 5-25 years.

| **N=106, obs=637** | **Coefficient (95% CI)** | **P-value** |
|---|---|---|
| Age | -1.61 (-2.06 – -1.15) | <0.001* |
| FIS | -4.11 (-10.64 – 2.43) | 0.218 |
| FIS*age | 0.49 (0.02 – 0.96) | 0.039* |
| Treatment | | |
| - none | Reference category | |
| - ivacaftor | 9.51 (4.85 – 14.18) | <0.001* |
| - lumacaftor/ivacaftor | -4.09 (-10.44 – 2.27) | 0.207 |
| Sex | | |
| - male | Reference category | |
| - female | 0.70 (-5.47 – 6.87) | 0.824 |
| Genotype class | | |
| - unclassified | Reference category | |
| - class I | -7.52 (-19.64 – 4.60) | 0.223 |
| - class II | -2.48 (-10.81 – 5.84) | 0.558 |
| - class III | 1.05 (-11.22 – 13.32) | 0.867 |
| - class IV | 11.85 (-3.47 – 27.16) | 0.129 |
| - class V | -21.71 (-41.16 - -2.26) | 0.029* |
| SCC | -0.09 (-0.24 – 0.06) | 0.241 |

*FEV1pp: Forced expiratory volume in 1 second, percent predicted. Age in years. FIS: Forskolin induced swelling, defined as the relative size increase of intestinal organoids (AUC) after 1h stimulation with 0.8 µM/L forskolin, AUC scaled 1:1000. FIS*age indicates the difference in annual FEV1pp decline per 1000 AUC change in FIS level. Genotype class: CFTR-protein function class of the mildest of both CFTR-mutations. SCC: Sweat chloride concentration in mmol/L.*

** significance level P < 0.05.*

**Supplementary table 4. Association of FIS with FEV1pp decline in sensitivity analysis including genotype group**

Regression coefficients linear mixed effects model FEV1pp with genotype group and subgroup only including participants with age between 5 – 25 years.

| | N=144, obs=1015 | | Subgroup: N=101, obs=605 | |
|---|---|---|---|---|
| | **Coefficient (95% CI)** | **P-value** | **Coefficient (95% CI)** | **P-value** |
| Age | -1.24 (-1.50 – -0.98) | <0.001* | -1.73 (-2.18 – -1.28) | <0.001* |
| FIS | -6.88 (-12.80 – -0.96) | 0.023* | -6.24 (-13.67 – 1.19) | 0.100 |
| FIS*age | 0.43 (0.22 – 0.63) | <0.001* | 0.54 (0.04 – 1.05) | 0.035* |
| Treatment | | | | |
| - none | Reference category | | Reference category | |
| - ivacaftor | 8.25 (4.62 – 11.88) | <0.001* | 10.21 (5.08 – 15.34) | <0.001* |
| - lumacaftor/ivacaftor | -3.97 (-8.41 – 0.47) | 0.079 | -4.31 (-10.65 – 2.02) | 0.182 |
| Sex | | | | |
| - male | Reference category | | Reference category | |
| - female | -0.44 (-6.38 – 5.51) | 0.886 | 0.82 (-5.63 – 7.26) | 0.804 |
| Genotype group | | | | |
| - Ins/del – missense | Reference category | | Reference category | |
| - Ins/del – nonsense | -1.47 (-11.04 – 8.10) | 0.763 | -0.03 (-9.96 – 9.89) | 0.995 |
| - Ins/del – splice | -5.92 (-15.31 – 3.47) | 0.216 | -5.03 (-15.33 – 5.27) | 0.337 |
| - Ins/del – ins/del | -2.50 (-11.75 – 6.76) | 0.597 | -0.08 (-10.17 – 10.02) | 0.988 |
| - Ins/del – unknown | 15.95 (-0.66 – 32.56) | 0.060 | 13.01 (-4.70 – 30.71) | 0.150 |
| - Missense – nonsense | 8.78 (-4.87 – 22.42) | 0.207 | 9.18 (-7.18 – 25.54) | 0.271 |
| - Missense – missense | 9.31 (-4.30 – 22.92) | 0.180 | 12.98 (-1.65 – 27.60) | 0.082 |
| - Splice – splice | -5.44 (-20.52 – 9.64) | 0.479 | 0.45 (-17.69 – 18.59) | 0.961 |
| SCC | -0.12 (-0.29 – 0.04) | 0.129 | -0.12 (-0.29 – 0.04) | 0.150 |

*FEV1pp: Forced expiratory volume in 1 second, percent predicted. Age in years. FIS: Forskolin induced swelling, defined as the relative size increase of intestinal organoids (AUC) after 1h stimulation with 0.8 µM/L forskolin, AUC scaled 1:1000. FIS*age indicates the difference in annual FEV1pp decline per 1000 AUC change in FIS level. Genotype group: combination of CFTR-mutation types on both alleles. SCC: Sweat chloride concentration in mmol/L.*

** significance level P < 0.05.*

**Supplementary table 5. Association of SCC with FEV1pp decline**

| **N=152, obs=1072** | **Coefficient (95% CI)** | **P-value** |
|---|---|---|
| Age | 0.25 (-0.86 – 1.37) | 0.654 |
| FIS | 1.71 (-2.97 – 6.38) | 0.473 |
| SCC | 0.07 (-0.16 – 0.31) | 0.544 |
| SCC*age | -0.01 (-0.02 – 0.00) | 0.064 |
| Treatment | | |
| - none | Reference category | |
| - ivacaftor | 9.35 (6.25 – 12.44) | <0.001* |
| - lumacaftor/ivacaftor | -0.68 (-4.80 – 3.44) | 0.747 |
| Sex | | |
| - male | Reference category | |
| - female | -1.18 (-7.79 – 5.44) | 0.727 |
| Genotype class | | |
| - unclassified | Reference category | |
| - class I | 1.43 (-11.77 – 14.63) | 0.832 |
| - class II | -1.47 (-10.24 – 7.31) | 0.743 |
| - class III | -2.42 (-15.30 – 10.46) | 0.712 |
| - class IV | -1.34 (-16.40 – 13.73) | 0.862 |
| - class V | -23.93 (-37.44 – -10.43) | <0.001 |

*FEV1pp: Forced expiratory volume in 1 second, percent predicted. Age in years. FIS: Forskolin induced swelling, defined as the relative size increase of intestinal organoids (AUC) after 1h stimulation with 0.8 µM/L forskolin, AUC scaled 1:1000. SCC: Sweat chloride concentration in mmol/L. SCC*age indicates the difference in annual FEV1pp decline per 1-unit change in SCC level. Genotype group: combination of CFTR-mutation types on both alleles. Genotype class: CFTR-protein function class of the mildest of both CFTR-mutations.*

** significance level P < 0.05.*

**Supplementary table 6. Comparison of the predictive value of FIS with SCC on FEV1pp decline**

| **N=152, obs=1072** | **Coefficient (95% CI)** | **P-value** |
|---|---|---|
| Age | -1.17 | 0.131 |
| FIS | -4.83 | 0.144 |
| FIS*age | 0.49 | 0.005* |
| SCC | -0.07 | 0.573 |
| SCC*age | -0.00 | 0.946 |
| Treatment | | |
| - none | Reference category | |
| - ivacaftor | 9.24 | <0.001* |
| - lumacaftor/ivacaftor | -0.60 | 0.774 |
| Sex | | |
| - male | Reference category | |
| - female | -1.39 | 0.678 |
| Genotype class | | |
| - unclassified | Reference category | |
| - class I | 0.73 | 0.913 |
| - class II | -2.19 | 0.623 |
| - class III | -2.93 | 0.653 |
| - class IV | -1.05 | 0.891 |
| - class V | -24.93 | <0.001* |

*FEV1pp: Forced expiratory volume in 1 second, percent predicted. Age in years. FIS: Forskolin induced swelling, defined as the relative size increase of intestinal organoids (AUC) after 1h stimulation with 0.8 µM/L forskolin, AUC scaled 1:1000. SCC: Sweat chloride concentration in mmol/L. FIS*age indicates the difference in annual FEV1pp decline per 1000 AUC change in FIS level. SCC*age indicates the difference in annual FEV1pp decline per 1-unit change in SCC level. Genotype group: combination of CFTR-mutation types on both alleles. Genotype class: CFTR-protein function class of the mildest of both CFTR-mutations.*

** significance level P < 0.05.*
